# 3D regional evaluation of right ventricular myocardial work from cineCT

**DOI:** 10.1101/2024.07.30.24311094

**Authors:** Amanda Craine, Anderson Scott, Dhruvi Desai, Seth Kligerman, Eric Adler, Nick H Kim, Laith Alshawabkeh, Francisco Contijoch

**Affiliations:** Department of Bioengineering, University of California San Diego, 9500 Gilman Drive, La Jolla, CA USA; Department of Radiology, University of California San Diego, 9500 Gilman Drive, La Jolla, CA USA; Division of Cardiovascular Medicine, University of California San Diego, 9500 Gilman Drive, La Jolla, CA USA; Division of Pulmonary, Critical Care, Sleep Medicine & Physiology, Department of Medicine, University of California San Diego, 9500 Gilman Drive, La Jolla, CA USA; Department of Radiology, National Jewish Health, 1400 Jackson Street, Denver, CO USA

## Abstract

**Background:** Regional myocardial work (MW) is not measured in the right ventricle (RV) due to a lack of high spatial resolution regional strain (RS) estimates throughout the ventricle. We present a cineCT-based approach to evaluate regional RV performance and demonstrate its ability to phenotype three complex populations: end-stage LV failure (HF), chronic thromboembolic pulmonary hypertension (CTEPH), and repaired tetralogy of Fallot (rTOF).

**Methods:** 49 patients (19 HF, 11 CTEPH, 19 rTOF) underwent cineCT and right heart catheterization (RHC). RS was estimated from full-cycle ECG-gated cineCT and combined with RHC pressure waveforms to create regional pressure-strain loops; endocardial MW was measured as the loop area. Detailed, 3D mapping of RS and MW enabled spatial visualization of strain and work strength, and phenotyping of patients.

**Results:** HF patients demonstrated more overall impaired strain and work compared to the CTEPH and rTOF cohorts. For example, the HF patients had more akinetic areas (median: 9%) than CTEPH (median: <1%, p=0.02) and rTOF (median: 1%, p<0.01) and performed more low work (median: 69%) than the rTOF cohort (median: 38%, p<0.01). The CTEPH cohort had more impairment in the septal wall; <1% of the free wall and 16% of the septal wall performed negative work. The rTOF cohort demonstrated a wide distribution of strain and work, ranging from hypokinetic to hyperkinetic strain and low to medium-high work. Impaired strain (−0.15≤RS) and negative work were strongly-to-very strongly correlated with RVEF (R=-0.89, p<0.01; R=-0.70, p<0.01 respectively), while impaired work (MW≤5 mmHg) was moderately correlated with RVEF (R=-0.53, p<0.01).

**Conclusions:** Regional RV MW maps can be derived from clinical CT and RHC studies and can provide patient-specific phenotyping of RV function in complex heart disease patients.

**Clinical Perspective:** Evaluating regional variations in right ventricular (RV) performance can be challenging, particularly in patients with significant impairments due to the need for 3D spatial coverage with high spatial resolution. ECG-gated cineCT can fully visualize the RV and be used to quantify regional strain with high spatial resolution. However, strain is influenced by loading conditions. Myocardial work (MW) – measured clinically derived as the ventricular pressure-strain loop area - is considered a more comprehensive metric due to its independence of preload and afterload. In this study, we sought to develop regional RV myocardial work (MW) assessments in 3D with high spatial resolution by combining cineCT-derived regional strain with RV pressure waveforms from right heart catheterization (RHC). We developed our method using data from three clinical cohorts who routinely undergo cineCT and RHC: patients in heart failure, patients with chronic thromboembolic pulmonary hypertension, and adults with repaired tetralogy of Fallot.

We demonstrate that regional strain and work provide different perspectives on RV performance. While strain can be used to evaluate apparent function, similar profiles of RV strain can lead to different MW estimates. Specifically, MW integrates apparent strain with measures of afterload, and timing information helps to account for dyssynchrony. As a result, CT-based assessment of RV MW appears to be a useful new metric for the care of patients with dysfunction.

## 1. Introduction

Right ventricular (RV) performance is a key prognostic indicator in various clinical populations including patients with heart failure, pulmonary hypertension, and congenital heart disease. However, profiling regional RV function is challenging for two reasons. First, the RV has a complex shape which limits 2D imaging, has a thin wall which necessitates high spatial resolution imaging, and can be difficult to fully capture with echocardiography due to its position in the chest. Second, strain metrics are sensitive to loading conditions (1,2). As a result, clinical assessments of RV function, such as 2D strain, have proven limited, especially in complex patient populations.

High-resolution, three-dimensional quantitative mapping of RV performance could improve understanding of RV pathophysiology and help inform pre-interventional evaluation. Contrast-enhanced, retrospectively-gated CT has been used to quantify regional strain (3–7). It is particularly well suited for RV analysis given imaging is inherently 3D and has high spatial resolution (<1 mm spatial resolution). Further, CT imaging can be performed in patients with implanted devices as well as very sick patients who can only tolerate short scan times. We have demonstrated how CT imaging of RV function improves evaluation of heart failure patients undergoing LVAD implantation (8–10). We have also used CT imaging to estimate regional strain throughout the RV and demonstrated that strain mapping can be used to capture regional dysfunction in adults with congenital heart disease (ACHD) (11).

However, strain metrics are influenced by preload and afterload. Specifically, strain mapping does not account for hemodynamic factors, such as pressure and volume overload. Regional myocardial work (MW), the area of the regional myocardial stress-fiber strain loop, has been proposed as a more robust measure of regional performance, as MW is independent of loading conditions and reflects regional oxygen perfusion and glucose metabolism (12,13). Clinically, global and regional MW has been measured via the ventricular pressure-strain loop area, which avoids challenges associated with quantifying myocardial stress (14,15).

In this study, we sought to map regional RV strain and MW by combining RV strain derived from ECG-gated cardiac CT with RV pressure recordings obtained from right heart catheterization (RHC). We also provide detailed phenotypes of RV performance. An overview of our approach is shown in **Figure 1**. We applied our approach to three distinct clinical populations who underwent CT imaging and RHC as part of their clinical care: patients with repaired tetralogy of Fallot (rTOF), patients with chronic thromboembolic pulmonary hypertension (CTEPH), and patients with left ventricular failure (HF). We hypothesize that 3D performance mapping will agree with the pathophysiology of each population and that MW mapping will complement strain-based evaluation. We observed that MW_CT_ enhances volumetric and strain-based assessments as highlighted by patients with differences in MW_CT_ despite similar RS_CT_ and volumetric findings.

**Figure 1:**
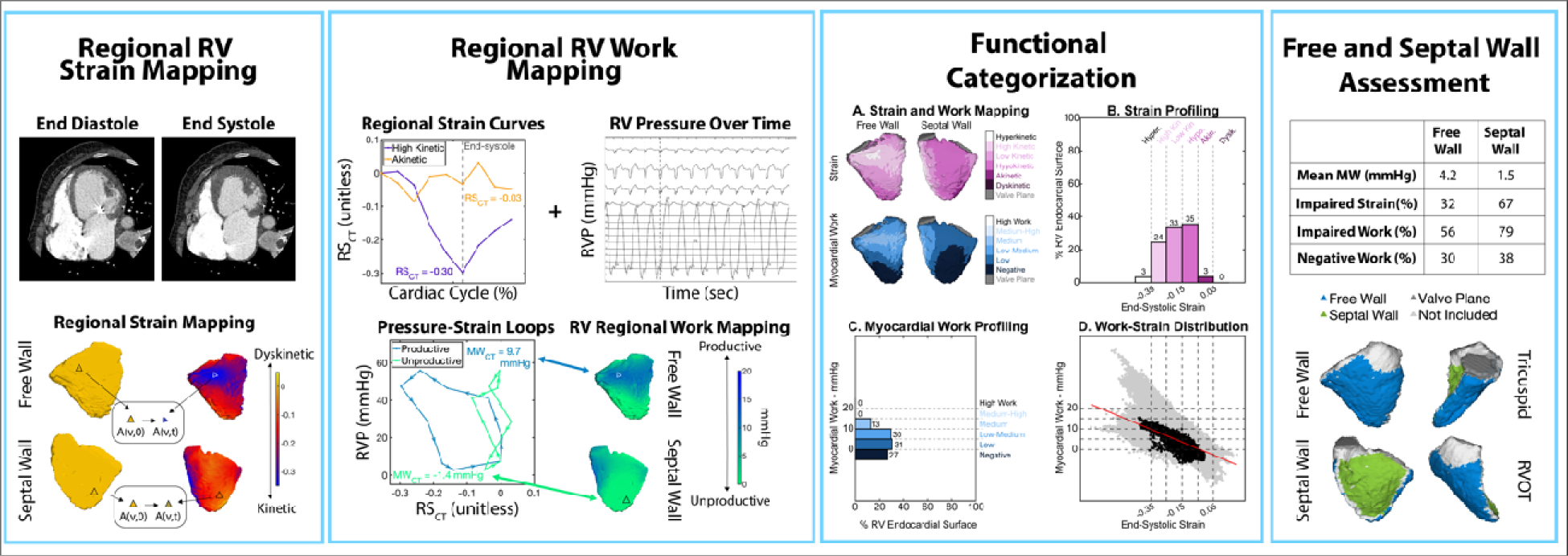
Categorization of RV function with regional work and strain. **Far Left**: Regional strain (RS_CT_) mapping via endocardial surface tracking with ECG-gated cineCT. 3D mapping of RS_CT_ highlights regional differences in end-systolic strain: blue/purple areas are kinetic while yellow/orange areas are dyskinetic. **Center Left:** RS_CT_ curves combined with RHC-derived RV pressure waveforms generate pressure-RS_CT_ loops. Myocardial work (MW_CT_) is estimated as the loop area. 3D mapping of MW_CT_ highlights regional differences in MW_CT_: blue indicates greater areas of work while green indicates areas performing less work. **Center Right:** RS_CT_ and MW_CT_ are separated into six categories to profile RV performance (B,C). RS_CT_ is categorized as dyskinetic (0.05≤RS_CT_), akinetic (−0.05≤RS_CT_<0.05), hypokinetic (−0.15≤RS_CT_<0.05), low kinetic (−0.25≤RS_CT_<-0.15), high kinetic (−0.35≤RS_CT_<-0.25), and hyperkinetic (RS_CT_<-0.35). MW_CT_ is categorized as negative (MW_CT_≤0 mmHg), low (0 mmHg<MW_CT_≤5 mmHg), low-medium (5 mmHg<MW_CT_≤10 mmHg), medium (10 mmHg<MW_CT_≤15 mmHg), medium-high (15 mmHg<MW_CT_≤20 mmHg), and high (20 mmHg <MW_CT_). 3D mapping shows the distribution of impairment across the RV. RV performance can also be analyzed in each wall. Sample patient is a 61–65 year-old with non-ischemic heart failure, NYHA FC 3, RVEDVI=104 mL/m2, RVSVI=46 mL/m2, RVEF=46%. The patient has significant impairments in both strain and myocardial work with performance being more limited in the septum than the free wall.

## 2. Methods

### 2.1 Patient population

Clinical records of adult patients (≥18 years) with either HF, CTEPH, or rTOF who underwent full cardiac cycle, ECG-gated contrast enhanced cardiac CT at our institution were reviewed to identify patients who also received contemporaneous RHC. CTEPH and HF patients underwent outpatient CT and RHC as part of their evaluation for pulmonary thromboendarterectomy (PTE) surgery or heart transplant/LVAD placement, respectively. Only patients who underwent RHC within 7 days of CT imaging were included. rTOF patients underwent CT and RHC as part of routine, outpatient evaluation, so patients who had no change to their cardiac status in between exams were included with no time restriction.

Patients were excluded if any cardiovascular event was documented between procedures. Patients with congenital heart disease diagnosis were excluded from the CTEPH and HF groups. Patients were not excluded due to weight, heart rate, or heart rhythm abnormalities. Patients were scanned according to current clinical protocols as discussed below. The Institutional Review Board (IRB) of University of California San Diego waived ethical approval of this work. Demographic and clinical data (sex, age, body mass index, previous medical history, functional class, relevant medications) were obtained from electronic patient records **(Table 1)**.

**Table 1:** Demographic and clinical patient information for HF, CTEPH, and rTOF patient cohorts. The rTOF cohort included more women, was expectedly younger, experienced a larger gap in time between receiving CT and RHC, had more severe PR and PS, and had better FC, RVSVI, RVEF, and CI. The CTEPH cohort had expectedly higher mPAP and PVR, and fewer instances of conductance disorders. The HF cohort had more instances of atrial fibrillation and pacemaker presence, higher HR, lower CO, and higher PCWP. Remaining patient history, CT and RHC measurements were comparable across groups. Units are reported next to each parameter. Median and interquartile ranges are presented unless otherwise indicated. Bold indicates significant differences across cohorts (p<0.05). Nearly all patients had New York Heart Association FC; 3 rTOF patients were classified with the World Health Organization scale. FC for one HF patient was labeled 3-4 respectively, and therefore rounded up. Valve dysfunction severity reported as in between labels (ex. mild/moderate) was rounded up. Of the valve function reported, 3 rTOF patients had PS rounded up, 6 rTOF and 1 HF patient had PR rounded up, and 3 rTOF and 1 HF patient had TR rounded up. BMI: body mass index. PS: pulmonary stenosis. PR: pulmonary regurgitation. TR: tricuspid regurgitation. FC: functional class. RVEDVI: RV end-diastolic volume index. RVESVI: RV end-systolic volume index. RVSVI: RV stroke volume index. RVEF: RV ejection fraction. HR: heart rate. CO: cardiac output. CI: cardiac index. mPAP: mean pulmonary artery pressure. RAP: right atrial pressure. PCWP: pulmonary capillary wedge pressure. PVR: pulmonary vascular resistance. *indicates normally distributed data

|  | HF (n = 19) | CTEPH (n = 11) | rTOF (n = 19) | p-value |
| --- | --- | --- | --- | --- |
| **Patient History** |  |  |  |  |
| Women, n (%) | 5 (26) | 6 (55) | 13 (68) | **0.03** |
| Age, years | 58 (51 - 69) | 57 (50 - 63) | 29 (26 - 33) | **<0.01** |
| BMI, kg/m<sup>2</sup> | 26.5 (22.7 - 29.0) | 28.9 (27.0 - 31.5) | 24.9 (23.0 - 29.5) | 0.32 |
| Age of TOF repair, months | N/A | N/A | 12 (8 - 26) | N/A |
| TOF repair with transannular patch, n (%) | N/A | N/A | 11 (58) | N/A |
| Prescribed riociguat, n (%) | N/A | 4 (36) | N/A | N/A |
| CTEPH disease level, n (%) |  | Left Right |  |  |
| 1 |  | 3 (27) 5 (45) |  |  |
| 2 | N/A | 4 (36) 4 (36) | N/A | N/A |
| 3 |  | 4 (36) 2 (18) |  |  |
| 4 |  | 0 0 |  |  |
| HF due to ischemia, n (%) | 6 (32) | N/A | N/A | N/A |
| HF receiving LVAD, n (%) | 6 (32) | N/A | N/A | N/A |
| Time between CT and RHC, days | 1 (0 - 4) | 2 (1 - 3) | 85 (36 - 146) | **<0.01** |
| Conductance disorders, n (%) | 19 (100) | 8 (73) | 18 (95) | **0.03** |
| History of atrial fibrillation, n (%) | 9 (47) | 0 | 2 (11) | **<0.01** |
| Pacemaker status, n (%) | 12 (63) | 3 (27) | 3 (16) | **0.01** |
| Presence of PS, n (%) |  |  |  |  |
| None | 19 (100) | 11 (100) | 3 (16) |  |
| Mild | 0 | 0 | 5 (26) | **<0.01** |
| Moderate | 0 | 0 | 8 (42) |  |
| Severe | 0 | 0 | 3 (16) |  |
| Presence of PR, n (%) |  |  |  |  |
| None | 4 (21) | 4 (36) | 1 (5) |  |
| Mild | 14 (74) | 7 (64) | 1 (5) | **<0.01** |
| Moderate | 1 (5) | 0 | 3 (16) |  |
| Severe | 0 | 0 | 14 (74) |  |
| Presence of TR, n (%) |  |  |  |  |
| None | 0 | 0 | 2 (11) |  |
| Mild | 13 (68) | 6 (55) | 12 (63) | 0.33 |
| Moderate | 5 (26) | 3 (27) | 4 (21) |  |
| Severe | 1 (5) | 2 (18) | 1 (5) |  |
| FC, n (%) |  |  |  |  |
| 1 | 0 | 1 (9) | 3 (16) |  |
| 2 | 0 | 3 (27) | 11 (58) | **<0.01** |
| 3 | 9 (47) | 6 (55) | 5 (26) |  |
| 4 | 10 (53) | 1 (9) | 0 |  |
| **CT Measurements** |  |  |  |  |
| RVEDVI, mL/m<sup>2</sup> | 102.8<br>(82.2 - 131.4) | 122.9<br>(106.3 - 170.7) | 118.6<br>(106.4 - 129.8) | 0.08 |
| RVESVI, mL/m<sup>2</sup> | 69.6<br>(49.6 - 88.8) | 79.8<br>(53.1 - 113.8) | 52.7<br>(42.6 - 64.0) | 0.05 |
| RVSVI, mL/m<sup>2</sup> | 34.0 (30.3 - 45.3) | 45.5 (40.5 - 60.9) | 63.3 (59.6 - 70.2) | **<0.01\*** |
| RVEF, % | 32.6 (31.1 - 43.1) | 39.9 (28.9 - 51.0) | 54.6 (50.9 - 58.3) | **<0.01** |
| **Catheterization Measurements** |  |  |  |  |
| HR, bpm | 92 (78 - 97) | 75 (74 - 85) | 74 (60 - 77) | **<0.01\*** |
| CO, L/min | 3.3 (2.6 - 4.2) | 3.9 (3.4 - 4.9) | 5.4 (4.4 - 6.7) | **<0.01\*** |
| CI, mL/min/m<sup>2</sup> | 1.8 (1.4 - 2.2) | 2.0 (1.7 - 2.4) | 3.0 (2.6 - 3.4) | **<0.01** |
| mPAP, mmHg | 31 (24 - 40) | 46 (36 - 51) | 21 (16 - 28) | **<0.01\*** |
| RAP, mmHg | 10 (6 - 11) | 12 (7 - 15) | 11 (9 - 15) | 0.13 |
| PCWP, mmHg | 22 (17 - 28) | 11 (11 - 13) | 13 (9 - 16) | **<0.01** |
| PVR, dynes s cm-5 | 198 (130 - 351) | 494 (457 - 870) | 87 (70 - 151) | **<0.01** |

### 2.2 CT Imaging

All patients underwent full cycle, ECG-gated cineCT imaging using a 256-slice Revolution CT scanner (GE Healthcare, Chicago, IL). Gantry rotation time was 280ms. As per the clinical protocol, patients were scanned at either 80, 100, or 120 kV based on BMI and the maximum tube current varied from 179-781 mA. Nearly all patient scans were reconstructed as 0.625 mm thick slices (n=1 HF, n=1 CTEPH, and n=2 rTOF scans were reconstructed at 1.25mm slices). Axial images were reconstructed at ∼10% intervals across the cardiac cycle (0 to 90% of the R-R interval) except for n=3 patients who had imaging from 0 to 80% of the R-R interval. Images were reconstructed on a 512×512 matrix using a standard reconstruction kernel. The field of view was typically 230 mm (range: 155 mm – 366 mm) which led to a typical in-plane resolution of 0.45 mm x 0.45 mm. RV volumetry (end-diastolic volume, end-systolic volume, stroke volume, ejection fraction) was obtained by segmenting the RV bloodpool at each time frame using an open-source region-growing algorithm (ITK-SNAP, Philadelphia, PA) (11,16).

### 2.3 Right ventricular pressure waveform acquisition

RHC hemodynamics values (heart rate, cardiac output, cardiac index, mean pulmonary artery pressure, right atrial pressure, pulmonary capillary wedge pressure, pulmonary vascular resistance) were obtained from electronic clinical reports. In addition, RV pressure waveforms were digitized alongside the simultaneously recorded ECG (WebPlotDigitizer). To synchronize the pressure waveform to the regional strain curves, a single R-R interval was selected.

### 2.4 Regional myocardial work estimation

Regional strain (RS_CT_) across the cardiac cycle was quantified for small triangular patches throughout the RV endocardial surface from the RV bloodpool segmentation (**Figure 1, left**) using MATLAB (MathWorks, Natick, MA), as described in Contijoch et al (11). Strain was synchronized with RV pressure to construct RV pressure-strain loops and estimate regional myocardial work (MW_CT_) (**Figure 1, center left**) (14,15). MW_CT_ and end-systolic (ES) RS_CT_ values were used to classify the performance of each patch of the RV endocardial surface. End-systolic RS_CT_ was defined as the minimum RS_CT_ value within 1 cardiac phase (∼10% cardiac cycle) of the minimum RV blood volume phase. Positive MW_CT_ describes work done *by* the patch of myocardium, while negative MW_CT_ reflects work done *to* the patch of myocardium (17).

3D mapping RV performance as MW_CT_ and end-systolic RS_CT_ (**Figure 1, center right**) shows spatial differences in the strength of work and strain. For quantitative phenotyping, RV end-systolic RS_CT_ was categorized as dyskinetic (0.05≤RS_CT_), akinetic (−0.05≤RS_CT_<0.05), hypokinetic (−0.15≤RS_CT_<-0.05), low kinetic (−0.25≤RS_CT_<-0.15), high kinetic (−0.35≤RS_CT_<-0.25), and hyperkinetic (RS_CT_<-0.35). RV MW_CT_ was categorized as negative (MW_CT_≤0 mmHg), low (0 mmHg<MW_CT_≤5 mmHg), low-medium (5 mmHg<MW_CT_≤10 mmHg), medium (10 mmHg<MW_CT_≤15 mmHg), medium-high (15 mmHg<MW_CT_≤20 mmHg), and high (20 mmHg<MW_CT_). Plotting MW_CT_ as a function of end-systolic RS_CT_ shows the relationship between work and strain. The spatial distribution of RV work and strain was visualized by a 3D color-coded rendering of the functional categories. We summarized regional MW_CT_ in the free wall (FW) and septal wall (SW) (**Figure 1, right**). The walls were manually delineated from the RV blood volume segmentations at end-diastole in ITK-SNAP.

**Figures 2-4** show representative examples for each cohort.

**Figure 2:**
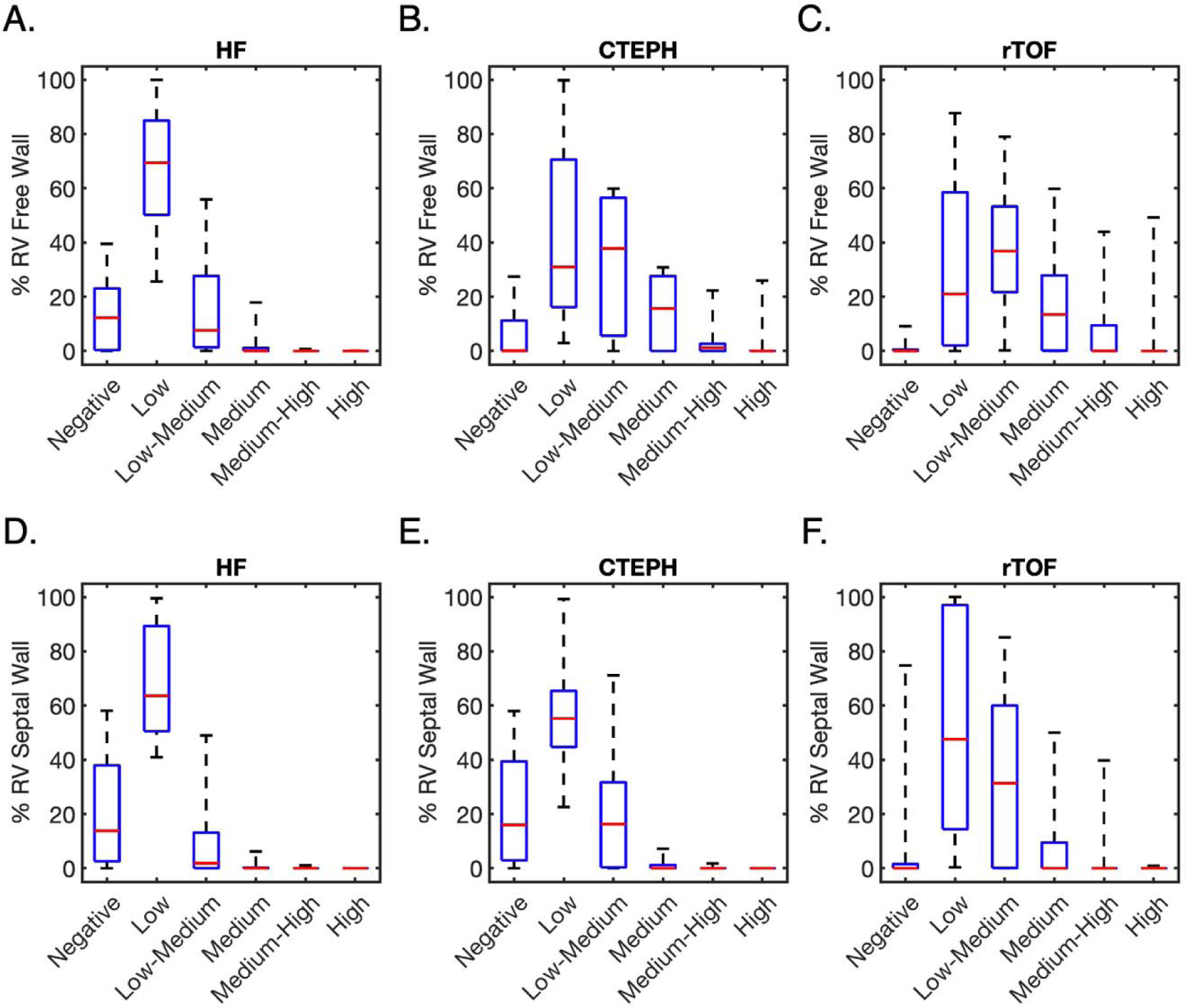
Evaluating regional RV MW_CT_ across cohorts. Categorization of myocardial work in the free wall and septal wall highlights differences between groups. In all groups, septal work was lower than free wall work. In the HF cohort, work performed by the two walls are very similar. In rTOF and CTEPH patients, free wall work values sometimes achieve medium-high or high levels of work but occurrences are more limited in the septal wall. In rTOF, we observed a large variation in the extent of negative work in the septal wall which is likely due to variations in surgical repair.

**Figure 3:**
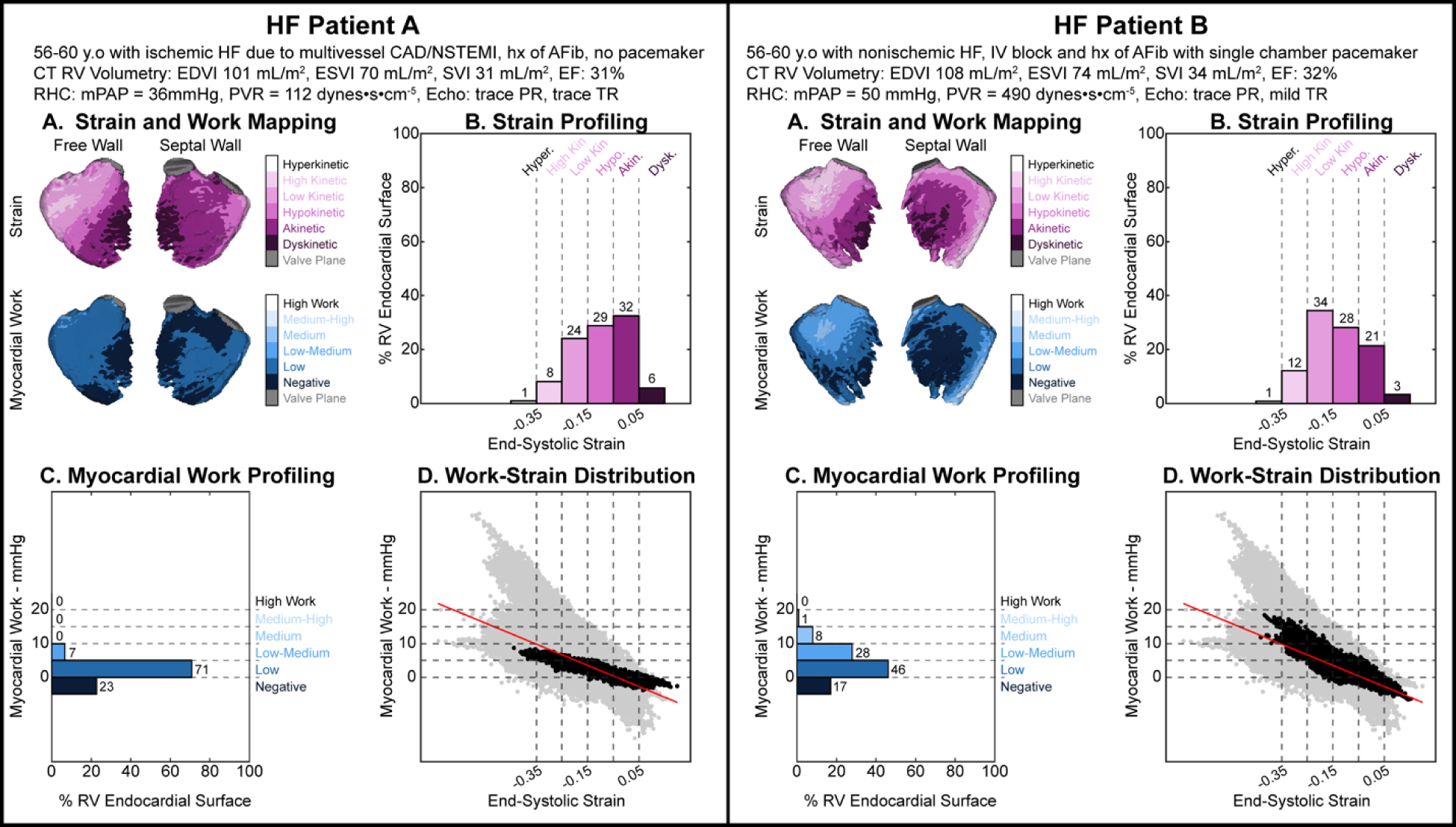
HF Cases with Similar Strain Profiles and Different Work Assessments. **Left:** HF patient A is a 56–60 year-old with ischemic heart failure due to multi-vessel CAD and NSTEMI, history of atrial fibrillation, no pacemaker, trace PR, trace TR, RVEDVI=101 ml/m^2^, RVESVI=70 ml/m^2^, RVSVI=31 ml/m^2^, RVEF=31%, mPAP=36 mmHg, PVR=112 dynes•cm^−5^•s. **Right:** HF patient B is a 56–60 year-old with nonischemic heart failure, intraventricular block, history of atrial fibrillation with a single chamber pacemaker, trace PR, mild TR, RVEDVI=108 ml/m^2^, RVESVI=74 ml/m^2^, RVSVI=34 ml/m^2^, RVEF=32%, mPAP=50 mmHg, and PVR=490 dynes•cm^−5^•s. **Strain profiling** (**Panels A and B**): HF patient A’s RV was 6% dyskinetic, 32% akinetic, 29% hypokinetic, 24% low kinetic, 8% high kinetic, and 1% hyperkinetic while patient B’s RV was 3% dyskinetic, 21% akinetic, 28% hypokinetic, 34% low kinetic, 12% high kinetic, and 1% hyperkinetic. In both patients, the RV free wall is dyskinetic and akinetic in the apex and along the RV insertion points on the RVOT side. Strain improves closer to the tricuspid valve plane, where the basal regions are mostly low kinetic and high kinetic. The RV septal wall is largely akinetic with dyskinetic areas in the mid region along the RV insertion points on the RVOT side. **Work Profiling (Panels A and C):** Patient A’s RV performed 23% negative work, 71% between 0–5 mmHg, and 7% between 5–10 mmHg. Patient B’s RV performed much better; 17% negative work, 46% between 0–5mmHg, 28% between 5–10 mmHg, 8% between 10–15 mmHg, and 1% between 15–20 mmHg. In particular, Patient B’s RV free wall performed much high work than patient A.

**Figure 4:**
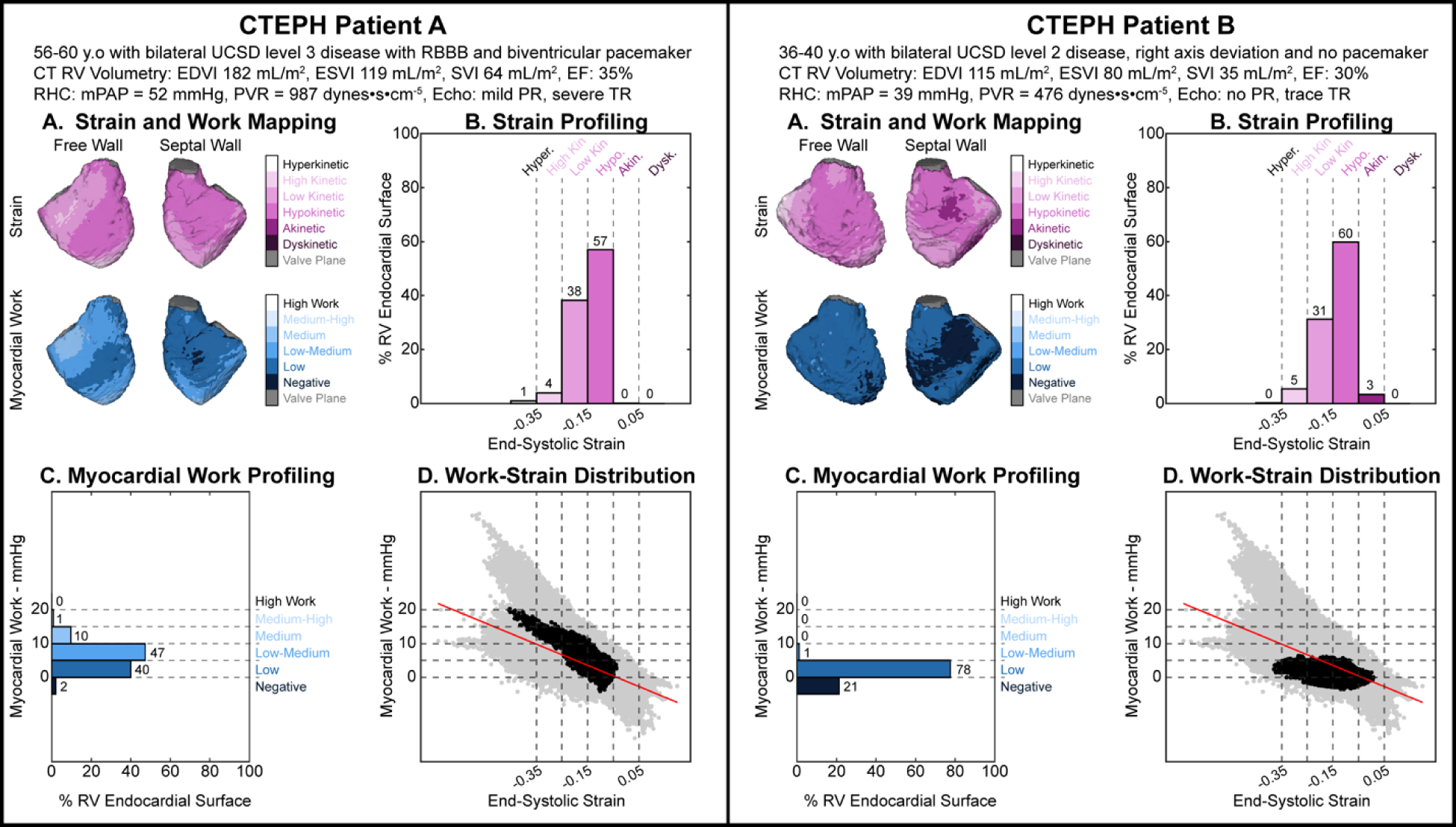
CTEPH Cases with Similar Strain Profiles and Different Work Assessments. **Left:** CTEPH patient A is a 56–60 year-old with bilateral UCSD level 3 disease, RBBB, and a biventricular pacemaker, mild PR, severe TR, RVEDVI=182 ml/m^2^, RVESVI=119 ml/m^2^, RVSVI=64 ml/m^2^, RVEF=35%, mPAP=52mmHg, and PVR=987 dynes·cm^−5^·s. **Right:** CTEPH patient B is a 36–40 year-old with bilateral UCSD level 2 disease, right axis deviation on ECG, no pacemaker, no PR, trace TR, RVEDVI=115 ml/m^2^, RVESVI=80 ml/m^2^, RVSVI=35 ml/m^2^, RVEF=30%, mPAP=39 mmHg, and PVR=476 dynes·cm^−5^·s. **Strain Profiling (Panels A and B):** Strain distribution between both patients was similar; Patient A’s RV was 0% akinetic, 57% hypokinetic, 38% low kinetic, 4% high kinetic, and 1% hyperkinetic while patient B’s RV was 3% akinetic, 60% hypokinetic, 31% low kinetic, and 5% high kinetic, and 0% hyperkinetic. In both cases, strain was impaired more in the septal wall than the free wall. **Work Profiling (Panels A and C):** Patient A’s RV performed much higher work than Patient B. Patient A’s RV performed 2% negative work, 40% between 0–5 mmHg, 47% between 5–10 mmHg, 10% between 10–15 mmHg, and 1% between 15–20 mmHg. Conversely, Patient B’s RV performed 21% negative work, 78% between 0–5 mmHg, and 1% between 5–10 mmHg.

### 2.5 Statistical analysis

For consistency, all results are presented as the median and interquartile range (Q1–Q3). Results were tested for normality via Shapiro-Wilk test. Results determined to be normal distributions are indicated with a starred p-value. Results determined to be non-normal distributions are analyzed with a non-parametric approach and the p-value is not starred. Results applicable to all three cohorts were evaluated via one-way ANOVA for normally distributed results and Kruskal-Wallis test for non-parametric results with a significance level of p<0.05. Agreement with non-normally distributed data was computed with Spearman correlation. When appropriate, post-hoc analysis was conducted to determine significance between groups. Indexed values were calculated with body surface area (BSA) via the Mosteller formula (18). Correlation coefficients were compared with Fisher r-to-z transformation.

## 3. Results

### 3.1 Patient History

The cohort of 49 patients comprised of 19 HF, 11 CTEPH, and 19 rTOF patients. 3/19 HF patients had RVEF>45%. Demographic and hemodynamic information is reported in **Table 1**. rTOF patients were younger (p<0.01), had longer time between RHC and CT (p<0.01), and had more severe pulmonary stenosis (PS, p<0.01) and pulmonary regurgitation (PR, p<0.01) compared to CTEPH and HF. The rTOF cohort included more women than the HF cohort (p=0.03). HF patients had more instances of conductance disorders than CTEPH (p=0.03), and more pacemakers than rTOF (p=0.01). HF patients had more instances of a history of atrial fibrillation (p<0.01) and worse functional class (FC) compared to CTEPH and rTOF (p<0.01). BMI and presence of tricuspid regurgitation (TR) were not significantly different amongst the groups.

### 3.2 CT-based Evaluation of RV Function

All groups had median RV end-diastolic volume index (RVEDVI)>100 mL/m^2^ (**Table 1**). Differences in RVEDVI and RV end-systolic volume index (RVESVI) were not significant but RV stroke volume index (RVSVI) was higher in the rTOF cohort (63 mL/m^2^, IQR: 60–70) than the HF cohort (34 mL/m^2^, IQR: 30–45, p<0.01) and the CTEPH cohort (46 mL/m^2^, IQR: 41–61, p=0.01). The CTEPH cohort also had greater RVSVI than the HF cohort (p=0.04). RV ejection fraction (RVEF) was higher in rTOF (55%, IQR: 51–58%) relative to CTEPH patients (40%, IQR: 29–51%, p<0.01) and HF patients (33%, IQR: 31–43%, p<0.01).

### 3.3 RHC Measurements

Heart rate in the rTOF cohort (74 bpm, IQR: 60–77) was lower than the HF cohort (92 bpm, IQR: 78–97, p<0.01). Cardiac output (CO) was higher in the rTOF cohort (5.4 L/min, IQR: 4.4– 6.7) than in CTEPH (3.9 L/min, IQR: 3.4–4.9, p=0.02) or HF (3.3 L/min, IQR: 2.6–4.2, p<0.01). Cardiac index (CI) was higher in the rTOF cohort (3.0 L/min/m^2^, IQR: 2.6–3.4) than the CTEPH cohort (2.0 L/min/m^2^, IQR: 1.7–2.4, p=0.02) and HF cohort (1.8 L/min/m^2^, IQR: 1.4–2.2, p<0.01). Mean pulmonary arterial pressure (mPAP) in the CTEPH cohort (46 mmHg, IQR: 36–51) was higher than rTOF (21 mmHg, IQR: 16–28, p<0.01) and HF (31 mmHg, IQR: 24–40, p=0.02). mPAP in the HF cohort was also greater than in rTOF (p=0.02). Pulmonary capillary wedge pressure (PCWP) was higher in the HF cohort (22 mmHg, IQR: 17–28) compared to rTOF (13 mmHg, IQR: 9–16, p=0.03) and CTEPH (11 mmHg, IQR: 11–13, p<0.01). Pulmonary vascular resistance (PVR) was higher in the CTEPH cohort (494 dynes s cm-5, IQR: 457–870) than the rTOF (87 dynes s cm-5, IQR: 70–151, p<0.01) and HF (198 dynes s cm-5, IQR: 130–351, p=0.01) cohorts. Right atrial pressure (RAP) was not different amongst the cohorts (p>0.05).

### 3.4 CT-based Quantification of RV Strain

RV performance was evaluated across patient populations (**Table 2**). The HF cohort had more akinetic areas (9%, IQR: 4–16) than the rTOF (1%, IQR: <1–4, p<0.01) and CTEPH cohorts (<1%, IQR: <1–5, p=0.02). The rTOF cohort had fewer hypokinetic areas (14% IQR: 9–18) than CTEPH (29%, IQR: 17–60%, p<0.01) and HF (35%, IQR: 29–48, p<0.01). The rTOF cohort also had more high kinetic (37%, IQR: 29–44) and hyperkinetic areas (10%, IQR: 7–16) than the CTEPH (high kinetic: 15%, IQR: 4–35, p=0.01; hyperkinetic: 2%, IQR: <1–4, p<0.01) and HF cohorts (high kinetic: 9%, IQR: 5–20, p<0.01; hyperkinetic: 1%, IQR: <1–5, p<0.01).

**Table 2:** RV Performance categorized by work and strain. The rTOF cohort performed the greatest amount of strain compared to the other groups, with most of the RV categorized as low kinetic and high kinetic. The CTEPH and HF patients were mostly hypokinetic and low kinetic. Despite limited RV strain, the CTEPH cohort had comparable MW to the TOF cohort, ranging from low to medium. The HF group performed less work, ranging from negative to low-medium. RV strain was categorized by the minimum RS_CT_ value at volumetric end-systole ± one cardiac phase. *indicates normally distributed data

|  | HF (n = 19) | CTEPH (n = 11) | rTOF (n = 19) | p-value |
| --- | --- | --- | --- | --- |
| **RV Strain Profiles** |  |  |  |  |
| Dyskinetic: $0.05 \leq RS_{CT}$, % | 0 (0 - 1) | 0 | 0 (0 - <1) | **0.05** |
| Akinetic: $-0.05 \leq RS_{CT} < 0.05$, % | 9 (4 - 16) | <1 (<1 - 5) | 1 (<1 - 4) | **<0.01** |
| Hypokinetic: $-0.15 \leq RS_{CT} < -0.05$, % | 35 (29 - 48) | 29 (17 - 60) | 14 (9 - 18) | **<0.01\*** |
| Low Kinetic: $-0.25 \leq RS_{CT} < -0.15$, % | 34 (24 - 44) | 39 (33 - 48) | 30 (25 - 41) | 0.09* |
| High Kinetic: $-0.35 \leq RS_{CT} < -0.25$, % | 9 (5 - 20) | 15 (4 - 35) | 37 (29 - 44) | **<0.01** |
| Hyperkinetic: $RS_{CT} < -0.35$, % | 1 (<1 - 5) | 2 (<1 - 4) | 10 (7 - 16) | **<0.01** |
| **RV Work Profiles** |  |  |  |  |
| Negative: $MW \leq 0$, % | 14 (1 - 24) | 5 (2 - 18) | 1 (<1 - 3) | **0.01** |
| Low: $0 < MW \leq 5$, % | 69 (56 - 82) | 40 (29 - 71) | 38 (18 - 75) | **0.01\*** |
| Low-Medium: $5 < MW \leq 10$, % | 9 (3 - 26) | 29 (6 - 47) | 29 (20 - 51) | **0.01** |
| Medium: $10 < MW \leq 15$, % | 0 (0 - 1) | 10 (<1 - 18) | 10 (<1 - 25) | **<0.01** |
| Medium-High: $15 < MW \leq 20$, % | 0 | 1 (0 - 2) | <1 (0 - 8) | **0.01** |
| High.: $20 < MW$, % | 0 | 0 (0 - <1) | 0 | 0.13 |

### 3.5 CT-based Quantification of RV Work

The HF cohort had more areas performing negative work (14%, IQR: 1–24) and low work (69%, IQR: 56–82) than the rTOF cohort (Negative: 1%, IQR: <1–3, p=0.01; Low: 38%, IQR: 18–75, p<0.01). The rTOF cohort had more areas performing low-medium work (29%, IQR: 20–51), medium work (10%, IQR: <1–25), and medium-high work (<1%, IQR: 0–8) than the HF cohort (Low-Medium: 9%, IQR: 3–26, p=0.01; Medium: 0%, IQR: 0–1, p<0.01; Medium-High: 0%, p=0.02).

### 3.6 Assessment of Regional RV Performance

Regional RV performance across patient populations is reported in **Table 3** and **Figure 2**. Metrics of RV strain and MW in the free wall were both higher than in the septal wall. Overall, trends observed in global assessment were also observed in the free wall. However, the rTOF cohort had fewer low kinetic areas in the RV free wall (27%, IQR: 13–35) than the CTEPH (38%, IQR: 36–51, p=0.01) cohort. In the septal wall, the HF cohort had fewer low kinetic areas (24%, IQR: 16–33) than rTOF (43%, IQR: 34–63, p<0.01) and CTEPH (39%, IQR: 31–57, p=0.01). Further, in the septal wall, the extent of negative work (rTOF: 0%, IQR: 0-2, CTEPH: 16%, IQR: 3-39, HR: 14%, IQR: 3-38) was significantly different amongst the groups, where the rTOF group had fewer areas of negative work than CTEPH (p=0.01) and HF (p<0.01).

**Table 3:** Regional RV performance categorized in the free wall and septal wall. Overall, the RV free wall performed greater work and strain then the RV septal wall, as expected. The HF cohort performed worse work and strain compared to the rTOF and CTEPH cohorts. The rTOF cohort performed higher RV free wall and septal wall strain than the CTEPHs and HF patients. RV free wall work was better in rTOF than in HF, but comparable to CTEPH. Negative work in the septal wall was lowest in the rTOF cohort. *indicates normally distributed data

|  | HF (n = 19) | CTEPH (n = 11) | rTOF (n = 19) | p-value |
| --- | --- | --- | --- | --- |
| **RV Free Wall Strain Profiles** |  |  |  |  |
| Dyskinetic, % | 0 (0 - <1) | 0 | 0 | **0.02** |
| Akinetic, % | 6 (<1 - 15) | <1 (0 - 2) | 0 | **<0.01** |
| Hypokinetic, % | 31 (26 - 52) | 20 (7 - 54) | 1 (<1 - 4) | **<0.01** |
| Low Kinetic, % | 38 (26 - 46) | 38 (36 - 51) | 27 (13 - 35) | **0.01\*** |
| High Kinetic, % | 13 (5 - 27) | 24 (5 - 45) | 52 (41 - 61) | **<0.01** |
| Hyperkinetic, % | 1 (<1 - 7) | 2 (<1 - 8) | 15 (10 - 26) | **<0.01** |
| **RV Free Wall Work Profiles** |  |  |  |  |
| MW ≤ 0, % | 12 (<1 - 23) | <1 (0 - 11) | 0 (0 - <1) | **<0.01** |
| 0 ≤ MW < 5, % | 69 (50 - 85) | 31 (16 - 71) | 21 (2 - 58) | **<0.01** |
| 5 ≤ MW < 10, % | 8 (1 - 28) | 38 (6 - 56) | 37 (22 - 53) | **0.01** |
| 10 ≤ MW < 15, % | 0 (0 - 1) | 16 (0 - 28) | 13 (<1 - 28) | **0.01** |
| 15 ≤ MW < 20, % | 0 | 1 (0 - 3) | <1 (0 - 9) | **0.01** |
| 20 < MW, % | 0 | 0 (0 - <1) | 0 | 0.23 |
| **RV Septal Wall Strain Profiles** |  |  |  |  |
| Dyskinetic, % | 0 (0 - <1) | 0 | 0 | 0.11 |
| Akinetic, % | 16 (7 - 32) | 1 (0 - 15) | 0 (0 - 2) | **<0.01** |
| Hypokinetic, % | 43 (31 - 59) | 38 (23 - 59) | 19 (11 - 23) | **<0.01\*** |
| Low Kinetic, % | 24 (16 - 33) | 39 (31 - 57) | 43 (34 - 63) | **<0.01\*** |
| High Kinetic, % | 2 (1 - 8) | 4 (<1 - 16) | 31 (14 - 39) | **<0.01** |
| Hyperkinetic, % | 0 (0 - 1) | 0 | 1 (0 - 7) | **0.03** |
| **RV Septal Wall Work Profiles** |  |  |  |  |
| MW ≤ 0, % | 14 (3 - 38) | 16 (3 - 39) | 0 (0 - 2) | **<0.01** |
| 0 ≤ MW < 5, % | 64 (51 - 89) | 55 (45 - 65) | 48 (14 - 97) | 0.28 |
| 5 ≤ MW < 10, % | 2 (<1 - 13) | 16 (<1 - 32) | 31 (<1 - 60) | 0.14 |
| 10 ≤ MW < 15, % | 0 (0 - <1) | 0 (0 - 1) | 0 (0 - 9) | 0.5 |
| 15 ≤ MW < 20, % | 0 | 0 | 0 | 0.86 |
| 20 < MW, % | 0 | 0 | 0 | 0.45 |

### 3.7 Profiling and 3D Mapping of RV Performance

**Figures 3** (HF), 4 (CTEPH), and 5 (rTOF) highlight assessment of RV strain and work in two patients from each cohort. Patients were selected based on similar strain profiles and clinical presentation. However, they present different work profiles.

In the heart failure cohort, we observed two patients with comparable RV volumetry and strain, but differences in work (**Figure 3**). Patient B had higher mPAP and PVR than patient A, which contributed to greater afterload. Furthermore, both patients had a history of atrial fibrillation, but only patient B had a pacemaker. Untreated atrial fibrillation appears to contribute to lower work values in patient A. Both patients received an LVAD, but only patient B developed post-operative RV failure.

In the CTEPH cohort, patient A had more variable work than patient B (**Figure 4**) despite similar strain patterns. Patient A had a much higher RVEDVI, PVR, and mPAP, indicating more severe pressure and volume overload. Therefore, the similar strain profiles observed led to patient A producing more work than patient B. For patient B, over 20% of the RV performed negative work. This could be attributed to RV dyssynchrony from untreated arrhythmia affecting strain since the patient had elevated mPAP and PVR.

In the rTOF cohort, patient A performed less work than patient B (**Figure 5**), despite a more homogenous work distribution. The higher work values observed in patient B were likely due to higher mPAP, given that the patients had comparable PS. The work heterogeneity observed between the free and septal walls in patient B was likely due to untreated right bundle branch block (RBBB). RBBB delays RV free wall depolarization, which would cause a mistiming between pressure and strain, resulting in altered work measurements.

**Figure 5:**
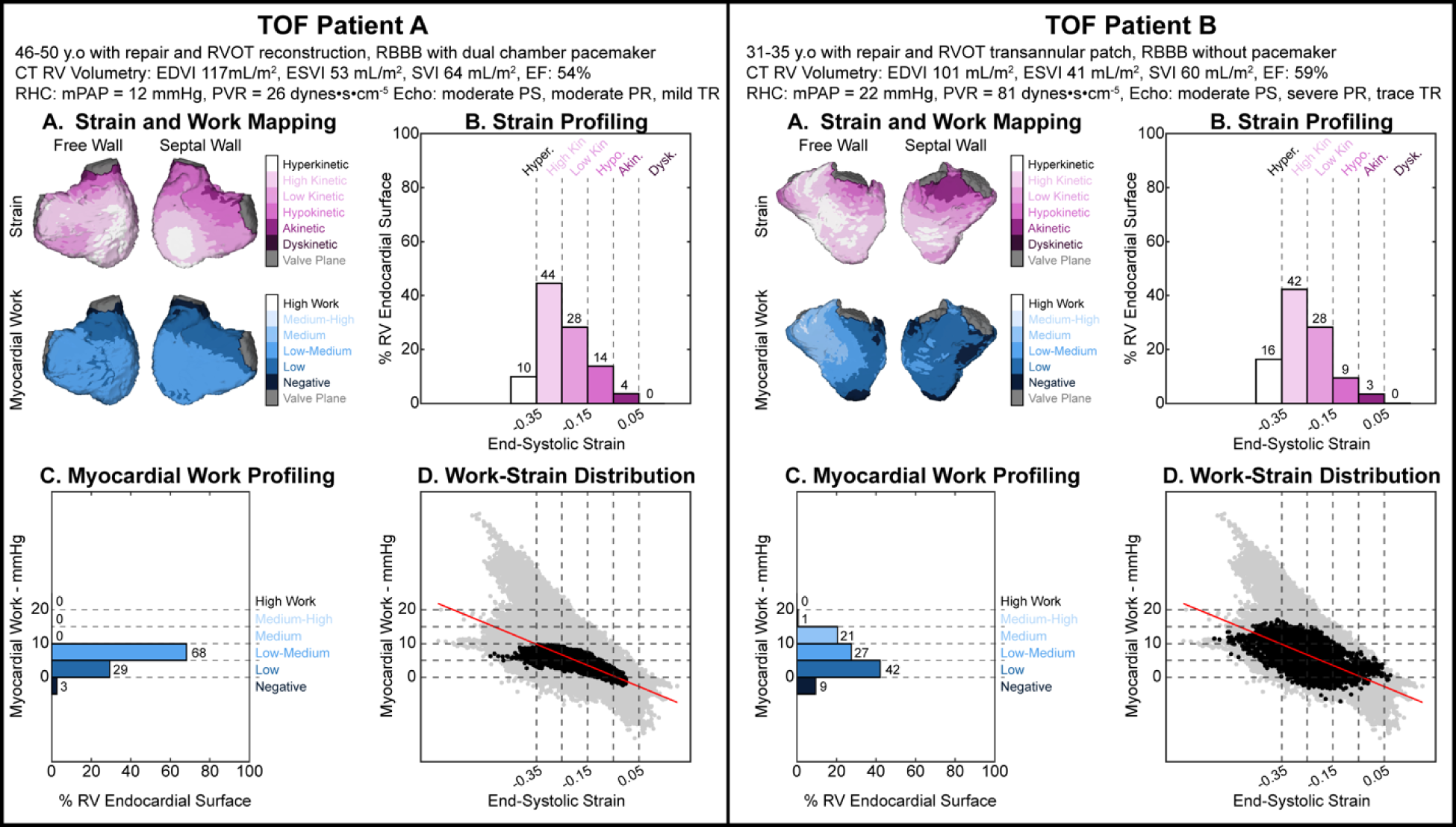
TOF Cases with Similar Strain and Diverging Work Assessment. **Left:** rTOF patient A is a 46–50 year-old with RVOT reconstruction, RBBB with a dual chamber pacemaker, moderate PS, moderate PR, mild TR, RVEDVI=117 ml/m^2^, RVEVI=53 ml/m^2^, RVSVI=64 ml/m^2^, RVEF=54%, mPAP=12 mmHg, and PVR=26 dynes•cm^−5^•s. **Right:** rTOF patient B is a 31–35 year-old with transannular patch, RBBB without a pacemaker, moderate PS, severe PR, trace TR, RVEDVI=101 ml/m^2^, RVEVI=41 ml/m^2^, RVSVI=60 ml/m^2^, RVEF=59%, mPAP=22 mmHg, and PVR=81 dynes•cm^−5^•s. **Strain Profiling (Panels A and B):** rTOF patients A and B have a similar distribution in their strain. Strain was mostly high kinetic and hyperkinetic in the free wall of both patients. In the septal wall, strain worsened from apex to base. Poorest strain occurred between the valves in both patients. **Work Profiling (Panels A and C):** rTOF patients A and B have different work profiles. Patient A’s RV mostly has MW_CT_ between 0–10 mmHg. MW_CT_ between 0–5 mmHg was localized to the RVOT region on the free wall and the basal region in the septal wall. Negative work was localized to the RVOT. Patient B had a more heterogeneous distribution of work. Negative work in the free wall was located in the apex and near the RVOT. The septal wall performed lower work than the free wall; the majority of the septal wall performed MW_CT_ between 0–5 mmHg, with patches of negative work and MW_CT_ between 5–10 mmHg.

### 3.8 Agreement between RV performance and Global Function

The association between the spatial extent of impaired RS_CT_ and MW_CT_ with RVEF is shown in **Figure 6**. The extent of impaired strain (defined as end-systolic −0.15≤RS_CT_) showed a very strong, negative correlation with RVEF (R=-0.89, p<0.01, **Figure 6A**). To evaluate different definitions of impaired work, we assessed the association between RVEF with the extent of negative work and impaired work (MW_CT_≤5 mmHg). In reference to our work categories, impaired work includes areas performing negative or low work. The extent of negative work showed a strong, negative correlation with RVEF (R=-0.70, p<0.01, **Figure 6B**) while the extent of impaired work showed a moderate, negative correlation RVEF (R=-0.53, p<0.01, **Figure 6D**). The correlation between the extent of impaired strain and RVEF was significantly higher than the correlation between negative work and RVEF (p=0.01) and the correlation between impaired work and RVEF (p<0.01). The correlation between the extent of negative work and RVEF was not significantly different from the correlation between the extent of impaired work and RVEF (p>0.05).

**Figure 6:**
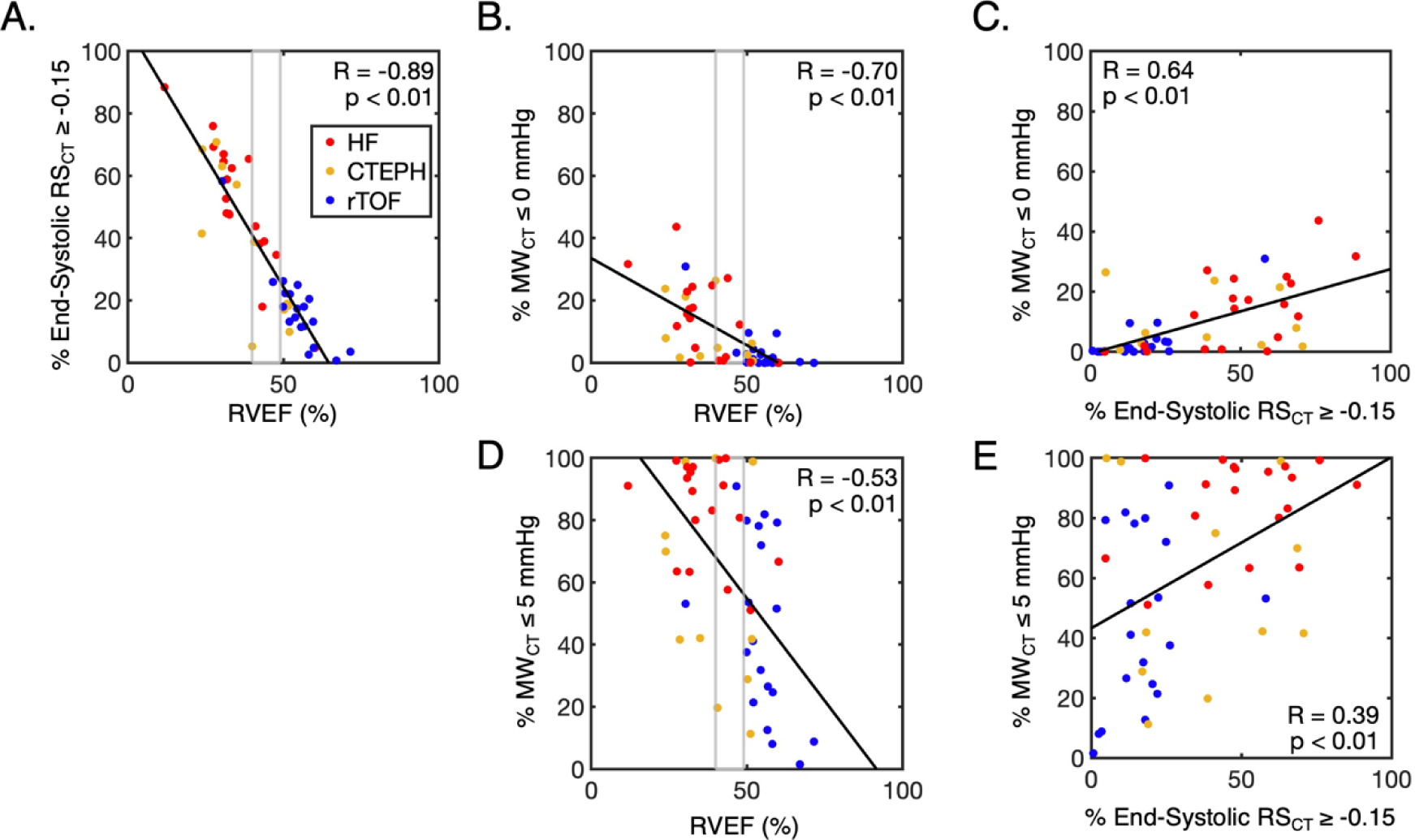
Agreement between impaired RV performance and global function. Data points representing HF patients are red, CTEPH patients are yellow, and rTOF patients are blue. Gray boxes indicate the extent of dysfunction in patients with RVEF between 40–49%. Agreement between the extent of impaired strain (−0.15≤RS_CT_) and RVEF is stronger than the extent of negative work or impaired work (MW_CT_≤5 mmHg) and RVEF. Agreement was determined with Spearman correlation.

Agreement between the spatial extent of impaired strain and negative work showed a strong, positive correlation (R=0.64, p<0.01, **Figure 6C**), while agreement between impaired strain and impaired work showed a weak, positive correlation (R=0.39, p<0.01, **Figure 6E**). These two correlations were not significantly different (p>0.05).

## 4. Discussion

In this study, we demonstrate a cineCT-based method to assess regional RV myocardial work with high spatial resolution throughout the endocardial surface and quantitatively phenotype patients. We estimated RV MW by combining clinically-derived RV pressure waveforms from RHC with 4D estimates of regional strain from ECG-gated, cardiac cineCT. We illustrated the utility of this method by profiling three clinical cohorts where both cineCT and RHC recordings are obtained as part of routine clinical care. By categorizing RV function with both myocardial work (MW_CT_) and strain (RS_CT_) measurements, we generated novel profiles of RV performance in each patient and for each cohort. Differences in 3D maps and quantitative metrics of RV performance agree with the clinical profile of our cohorts.

CineCT evaluation of regional strain has focused primarily on end-systolic strain. However, the ability of cineCT-based metrics to evaluate temporal differences in strain is of clinical interest (19). Motion artifacts associated with single-beat CT imaging can falsely identify dyssynchrony in healthy patients (19,20). By accounting for differences in loading conditions and integrating strain estimates across the cardiac cycle, MW offers the possibility of distinguishing areas of dyssynchrony from areas of dyskinesia without advanced correction approaches (20).

### 4.1 RV performance analysis with MW_CT_ and RS_CT_

We demonstrate how MW_CT_ complements volumetric evaluation and RS_CT_ mapping by highlighting patients with differences in MW_CT_ despite similar RS_CT_ findings. We believe MW_CT_ provides multiple advantages over strain metrics, as the pressure waveform impacts work estimates in two key ways. First, the amplitude of the pressure waveform accounts for afterload variations and allows for quantitative assessment across patients. In our study, this was critical for accurate assessment of patients with PS (many of the rTOF patients) as well as patients with PH (the CTEPH cohort). The shape of the pressure waveform is also important as it provides global timing information, enabling MW_CT_ to identify regions of inefficient endocardial motion (i.e dyssynchrony). The rTOF and HF patients had high prevalence of conductance disorders which would be incompletely evaluated using RS_CT_ alone.

Another benefit of our regional approach is that it enables evaluation of both the spatial extent and severity of mechanical and energetic impairment. Our 1D (**Figure 3-5BC**), 2D, and 3D approaches to mapping RV function highlight the value in measuring several complementary metrics of function. End-systolic RS_CT_ captures function at one time point whereas MW_CT_ evaluates performance throughout the entire cardiac cycle. 3D mapping (**Figure 3A-5A**) highlights spatial differences in work and strain within clinical populations. We observed discrepancies in work profiles between strain profile pairs, and across clinical populations, as demonstrated by clinical profiles matching trends in work and strain profiles. However, our study did not have longitudinal imaging, so it remains unclear how the distribution of these strain and work profiles change over time.

Global RV MW has been previously evaluated by Butcher et al with 2D echocardiography (14). By using 3D cineCT, we evaluated the entire RV endocardial surface, quantified the spatial extent of impairment, and analyzed regional RV performance. Despite methodological differences, our findings were largely consistent with Butcher (14), which supports the notion of combining work and strain measurements to provide a more detailed description of RV function globally and regionally by considering loading conditions, dyssynchrony, and function over the whole cardiac cycle.

In these studies, global RV MW (measured as global work index: GWI) was measured in normal cohorts (14,21) using global longitudinal strain curves. In our study, we developed MW categories based on values we observed, and not those described earlier (14,21,22), for several reasons. First, our study did not include normal patients, so “normal” values for our method have not been characterized. Further, our method differs from global longitudinal strain mapping in that a patch of the endocardium is evaluated, and strain is measured in a non-directional manner. Lastly, it is unclear whether global MW values like GWI should be applied to individual patches of the RV. Additional study is needed to determine normal ranges for regional RV MW in a population without cardiovascular disease and identify normal variations/patterns of RV MW throughout the RV surface.

### 4.2 Clinical Observations in RV Performance

Our approach to myocardial work profiling highlighted differences in RV performance across three clinical cohorts. Our findings agreed with their clinical profile. Specifically, our HF cohort had elevated PCWPs compared to the other groups, 79% had pulmonary hypertension (mPAP>20 mmHg) (23), and 84% had RV dysfunction (RVEF≤45%) (24). Amongst the RV segments, MW_CT_ was comparably reduced in the FW and SW; septal impairment is expected due to the presence of LV failure, and FW impairment is consistent with maladaptive remodeling. Further, MW_CT_ across the RV and FW was significantly lower in the HF cohort. This is consistent with a low output state. HF patients who receive an LVAD are at risk of RV failure after implantation (25). RV size and stroke work index (SWI) have been shown to predict post-LVAD RV failure (8,9). However, the HF patient pairs shown in Figure 3 had similar RV volumetry but different outcomes post-LVAD implantation. Previous literature has demonstrated that the difference in pre-operative free wall and septal wall strain is predictive of RV failure after LVAD implantation (10). Therefore, MW_CT_ may be useful in stratifying risk in HF patients seeking advanced therapies. However, the prognostic value of regional myocardial work should be evaluated in a dedicated study with longitudinal follow up of patients.

The CTEPH group had more impaired RV strain than patients with rTOF. We attribute this to increased afterload (26). This is supported by hemodynamics findings, such as elevated mPAP and PVR (26). Amongst the RV segments, impaired strain and work was common in the septal wall, which suggests impingement on the LV (27). Septal bowing is observed clinically and is known to compromise septal shortening (28). As the septum bows into the LV in response to increased RV pressure, the septum fibers rotate from an oblique position to a transverse position (28), which has been shown to delay RV systole (29) and limit the septum’s ability to shorten (28,30). Butcher et al showed that reduced RV work was associated with mortality in patients with PH (21). Future work should evaluate whether a regional metric which can separate the RV free wall from the septum improves this association. Further, whether MW_CT_ improves selection of patients for therapies or interventions is left for future study.

rTOF patients had more heterogeneous distributions of MW_CT_ in the RV free wall than the septal wall. Limited MW_CT_ in the RV septal wall is consistent with ventricular septal defect repair; late gadolinium enhancement (LGE) is observed at the site of septal defect patching in adults with rTOF (31). Heterogeneity in free wall MW_CT_ was attributed to the wide range of valvular dysfunction observed in this cohort. As a result of surgical repair of the RVOT, TOF patients often develop pulmonary regurgitation (n=18/19 in our cohort), which leads to volume overload (32). Repair can leave residual RVOT obstruction leading to pulmonary stenosis (n=16/19) and increased afterload (33). This mixed pulmonary disease is known to cause abnormal remodeling (34), and we attribute the combination of pressure and volume overload observed in varying degrees to the wide range of work values observed in this cohort.

3D mapping of RV strain and work demonstrates that patients with the same primary diagnosis, similar volumetric evaluation, and similar strain profiles, can have different work profiles due to differences in dyssynchrony and hemodynamics. Therefore, work profiling can inform several factors about a patient’s cardiac status summarized into one quantitative map.

### 4.4 Agreement between MW_CT_ and RS_CT_ and global function

Agreement between the extent of impaired strain (−0.15≤RS_CT_) and RVEF (**Figure 6A**) was stronger than agreement between the extent of negative work (**Figure 6B**) or impaired work (MW_CT_≤5 mmHg) and RVEF (**Figure 6D**). This observation supports that MW_CT_ and RS_CT_ provide different functional information. Specifically, patients with RVEF 40-49% (**Figure 6**, gray boxes) can have a wide range of tissue performing impaired work. However, such a relationship was not observed between RVEF and areas of impaired strain. This may be attributed to MW_CT_ incorporating function over the whole cardiac cycle, while dyskinesia is a characteristic of RS_CT_ only at end-systole. Ultimately, this observation suggests that MW_CT_ may detect aspects of RV dysfunction that cannot be identified by strain or RVEF, which could aid diagnosis or improve treatment planning.

### 4.5 Limitations

This study has several limitations. First, we did not analyze normal patients. While cardiac CT scans may be obtained in patients who end up having normal structure and function, such a finding would typically prevent a patient from undergoing RHC. As a result, for this retrospective study, we chose to evaluate patients with three different clinically diagnosed impairments to showcase the ability of MW_CT_ to identify spatial differences in dysfunction. Second, patients did not undergo RHC and CT imaging simultaneously. However, recent work in our group has shown that combining non-simultaneous RV pressure with CT volumes to estimate stroke work index in our HF cohort compares favorably to clinical estimates (9). Third, we utilized the pressure waveform as a surrogate for myocardial stress. This simplification ignores the spatial heterogeneities in myocardial stress that can arise due to chamber geometry and differences in material properties. An advantage to this approach is that it only requires clinical data that is routinely collected and has been demonstrated to be useful in LV and RV evaluation in prior studies (12,14,15,35,36). However, these studies have utilized cardiac magnetic resonance (12) and echocardiography (14,15,36) to acquire ventricular strain. Fourth, as a retrospective analysis of patients with CT images, we did not compare MW_CT_ to other imaging modalities such as MRI. However, prior work has shown agreement between our CT strain estimates and CMR tagging (6,7). This is a single-center study which should be confirmed in other cohorts, and we did not evaluate the prognostic value of MW_CT_ mapping. Lastly, this study included a relatively small number of cases from three clinical cohorts. Larger studies are needed to confirm our findings and implement our approach for clinical use.

## Data Availability

Research code and deidentified patient data reported in this article will be made available upon acceptance for publication in a Github repository.

## Acknowledgements

None

